# A qualitative examination into the support needs of people living with obesity during transition from tertiary obesity treatment

**DOI:** 10.1101/2022.03.19.22271723

**Authors:** Ghada Alsultany, Aymen El Masri, Freya MacMillan, Kathryn Williams, Kate McBride

**Affiliations:** School of Medicine, Western Sydney University, Penrith, NSW, 2751, Australia; School of Health Sciences, Western Sydney University, Penrith, NSW, 2751, Australia; Translational Health Research Institute (THRI), Western Sydney University, Penrith, NSW, 2751, Australia; Charles Perkins Centre–Nepean, Faculty of Medicine and Health, The University of Sydney, Kingswood, New South Wales, Australia; Department of Endocrinology, Nepean Hospital, Nepean Blue Mountains Local Health District, Kingswood, New South Wales, Australia

**Author notes:** **Corresponding author:** Dr Kate McBride. **Declarations of interest:** None to declare.

**Keywords:** Obesity, Transition, Community care, Integrated care, General practitioners

## Abstract

**Background:** As the number of people living with obesity increases, the maintenance of treatment outcomes is especially pertinent. Treatment at tertiary obesity services have proven to be successful, but patients need to be transitioned out of these services to community-based care to accommodate the influx of new patients. Little is known about the support needs of patients after transition from acute tertiary obesity services. It is important to establish the supports needed by these patients, especially in the context of maintaining treatment outcomes and ensuring continuity of care.

**Methods:** A qualitative study was conducted to identify the support needs of people with obesity as they transition to community care. Patients and clinicians recruited from a tertiary obesity clinic participated in semi-structured interviews and focus groups to explore factors influencing transition and supports needed in the community. Data was collected through audio recordings, transcribed verbatim and analysed thematically.

**Results:** A total of 16 patients and 7 clinicians involved in the care of these patients participated between July 2020 and July 2021. Themes identified included the influence of clinic and individual factors on transition, the benefits of phased transition, patient-centred communication, and the role of social support. It was found that dependency and lack of self-efficacy, as well as low social support, hindered transition efforts. It was also identified that patients required substantial integrated professional and social support structures in the community to adequately address their care needs both during and following transition.

**Conclusion:** Interventions are needed to provide social community services following transition to ensure adequate community care that can support the maintenance of treatment outcomes. Such services should be integrated and address the social needs of people living with obesity.

## 1. Introduction

Over 65% of adults and 25% of children in Australia are overweight or living with obesity and, as a result, live with a range of complications such as diabetes and cardiovascular disease[1]. The World Health Organization (WHO) classifies obesity as a chronic condition resulting from excessive fat accumulation increasing risks to physical and mental health, as well as heightening social and economic burdens on both individuals and communities[2]. For simplicity, obesity is measured as a body mass index (BMI) of ≥30 kg/m^2^ or more.[1].

In Australia, severe cases of obesity are treated at tertiary level, often hospital-based, services, which have proven successful in achieving treatment outcomes, such as weight loss, improved metabolic parameters, and behavioural and psychological skill building[3]. Patients are often transitioned out of these services into community-based care following the acute phase of treatment, to accommodate the influx of new patients[4]. Transition out of hospital services is a crucial stage, as maintenance of treatment outcomes is required for sustainability of weight loss.

Adequate services are needed to support patients as they transition out of obesity services to community care, as acute clinic interventions are unlikely to have continued efficacy without long term community support due to the chronic and often lifelong nature of obesity[2, 4]. While primary care and allied health services exist, there may be issues in the integration of these services with tertiary care[3]. The care needs of people with obesity are often unique, requiring specialised equipment and discharge planning when transitioning from hospital to home[5]. As such, it is important to explore the supports and services needed during transition out of tertiary obesity clinics for people with obesity, as this phenomenon has not previously been sufficiently examined through qualitative techniques.

Patients’ perspectives on their own care can bring important insights on how care should be delivered, ensuring client-centred services that appropriately address their needs[6]. In addition, exploring and comparing the perspectives of clinicians with patients can optimise and improve services, while addressing potential barriers to care[7].

As such, we aimed to explore the supports needed by patients during transition from a tertiary obesity clinic in an Australian healthcare setting through a qualitative exploration of the perspectives of both patients and clinicians.

## 2 Methods

Exploring patients and clinicians perspectives was achieved through a qualitative study employing semi-structured interviews and focus groups, described in accordance with the consolidated criteria for reporting qualitative studies (COREQ) checklist[8]. Ethics approval was granted by the Nepean Blue Mountains Human Research Ethics Committee 2019/ETH13681.

### 2.1 Participant recruitment

Purposive sampling was used to recruit participants. Participants were recruited from a public tertiary level hospital-based obesity service based in a local health district in Greater Western Sydney. The area within which the service is located has the highest rate of obesity (66.9%) across all metropolitan Primary Health Networks (PHNs) in Australia[1].Current or previous adult patients, and clinicians involved in their care, were invited to participate in the study. Patients were recruited through flyers and referrals from clinic staff and their peers, while clinicians were contacted by a researcher via email and invited to participate in one of two focus groups. Informed consent was obtained in writing or online, depending on the mode of referral and participant preferences.

### 2.2 Data collection

Participants were contacted by phone or email to schedule interviews and focus groups which were conducted through an online platform (Zoom), by phone, or face-to-face either at the service or a convenient location. Data were collected using semi-structured focus groups and one-on-one interviews between December 2020 and July 2021. The interview guide was developed based on existing literature, research experience, and expert contribution. Questions focused on identifying existing support and further support needed from professional and social networks during transition.

Interviews lasted between 30 – 60 minutes and were recorded using a digital voice recorder before being transcribed verbatim using secure third-party services, with all identifying information removed from the transcripts. Only researchers and participants were present during data collection. No repeat interviews were conducted, and transcripts were not returned to participants. Data collection was ceased when data saturation was determined, as no new major themes arose from the data[9].

### 2.3 Data analysis

Data was thematically analysed using an inductive approach, as outlined by Braun and Clarke[10]. De-identified transcripts were coded using the Quirkos program (Quirkos, Edinburgh, UK). Transcripts were checked for accuracy and re-read concurrent to code and theme generation to facilitate data familiarisation. Initial codes were generated inductively across the entire data set and initial themes were developed by finding patterns across these codes. Themes were reviewed and refined then defined and named in terms of the scope and focus of each theme, resulting in a narrative analysis reporting themes and data excerpts in tabular form. Participant identification numbers and pseudonyms were used to label excerpts. The coding framework was independently established by GA and KM then compared, discussed, and 25% of the total data were member checked by KW and AE to promote trustworthiness of the data interpretation.

### 2.4 Research team/reflexivity

The research team consisted of active University faculty members, clinicians and a research student. All have had experience or training in qualitative research methods. Participants were interviewed by KM and GA. No previous relationship existed between researchers and participants and participants were made aware of the credentials of the interviewers and motivations for the research. The research team were aware of assumptions and biases they may have had and addressed these through group discussion of results to promote reflexivity.

## 3. Results

A total of twenty-three patients and seven clinicians were contacted. Seven patients did not participate due to inability to make contact, leaving sixteen patients (13 female) and seven clinicians who were interviewed and included in this study.

Thematic analysis resulted in three main categories being identified; service-related factors influencing transition, individual-related factors influencing transition, and bridging the gap to facilitate transition.

### 3.1 Service-related factors influencing transition

Service-related factors influencing transition included the role of the clinic and how staff facilitated transition. This involved organisation of the transition process, influence of both positive and negative clinic communication, as well as patients feeling cared for. Clinic resources and services also influenced transition, though constraints of funding hindered their provision (Table 2).

**Table 1:** Demographic data for participants

| Patient characteristics | Mean (range)/count (%) |
| --- | --- |
| **Age** | 51 (35-65) |
| **Gender** |  |
| **Female** | 13 (81%) |
| **Male** | 3 (19%) |
| **Clinic patient** |  |
| **Current** | 9 (56%) |
| **Past** | 7 (44%) |
| **Part of social support group** |  |
| **Yes** | 8 (50%) |
| **No** | 8 (50%) |
| **Clinician characteristics** |  |
| **Gender** |  |
| **Female** | 6 (86%) |
| **Male** | 1 (14%) |
| **Role** |  |
| **Clinical Psychologist** | 2 |
| **Dietician** | 3 |
| **Endocrinologist** | 1 |
| **Physiotherapist** | 1 |

**Table 2:**
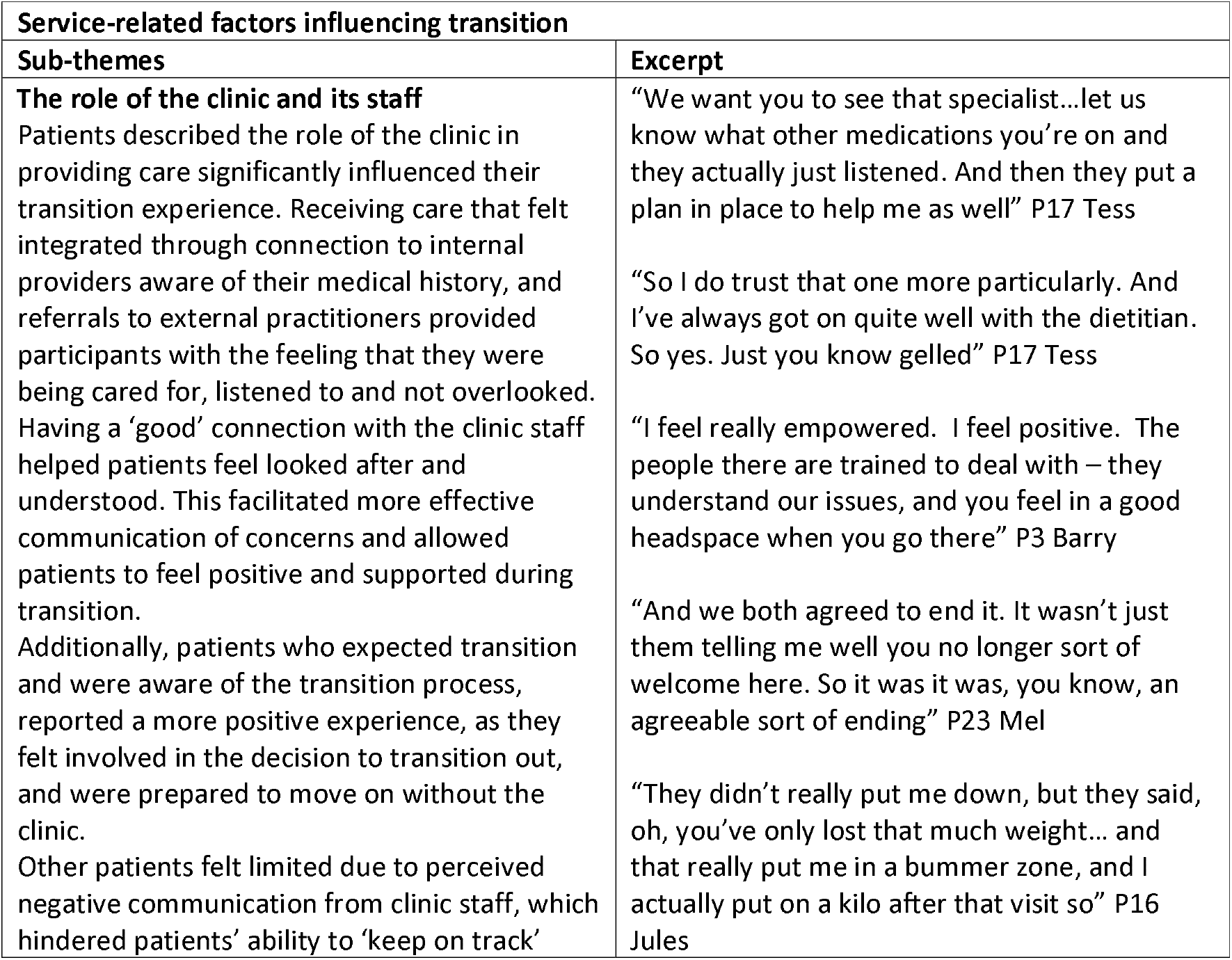
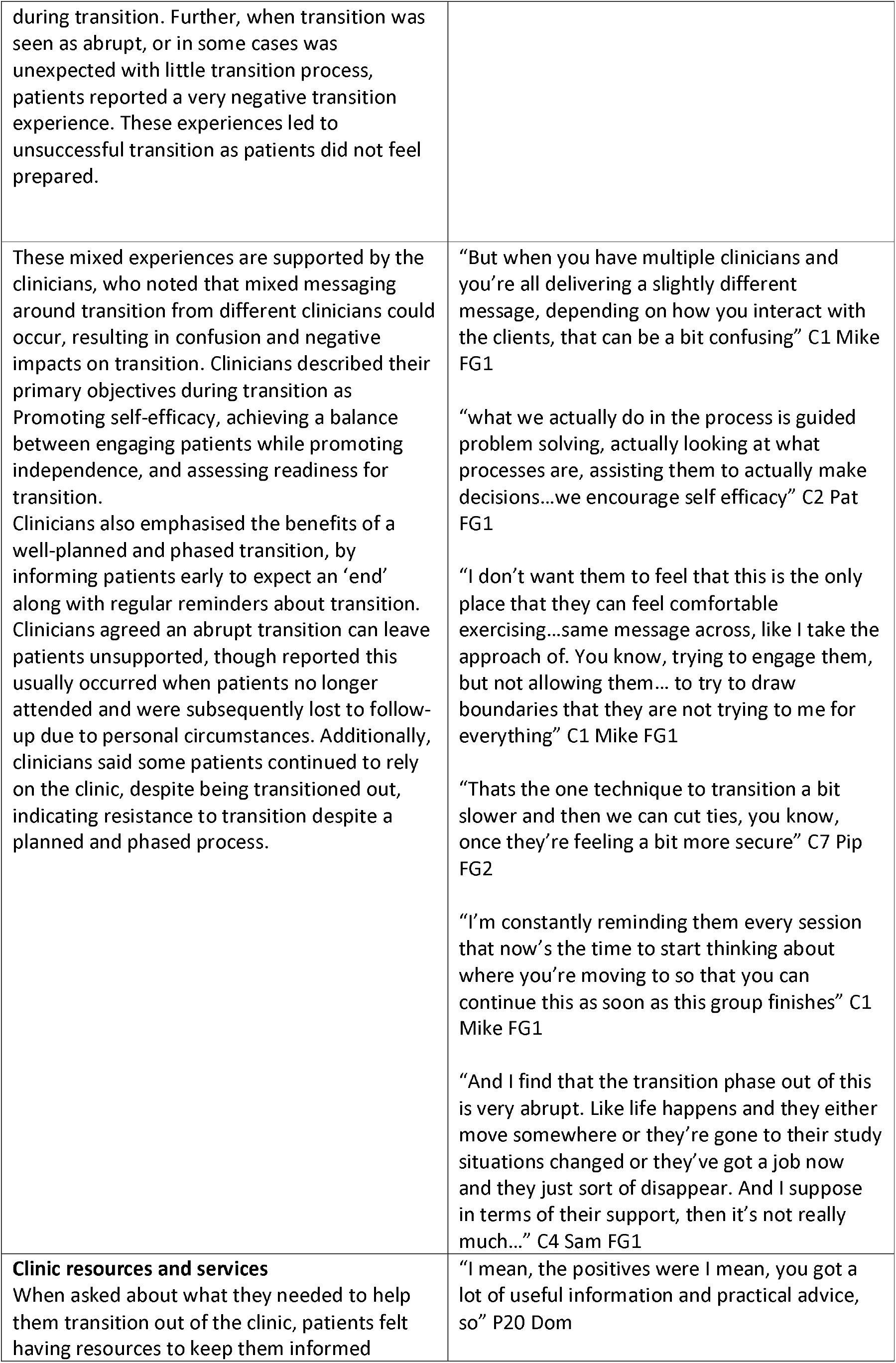
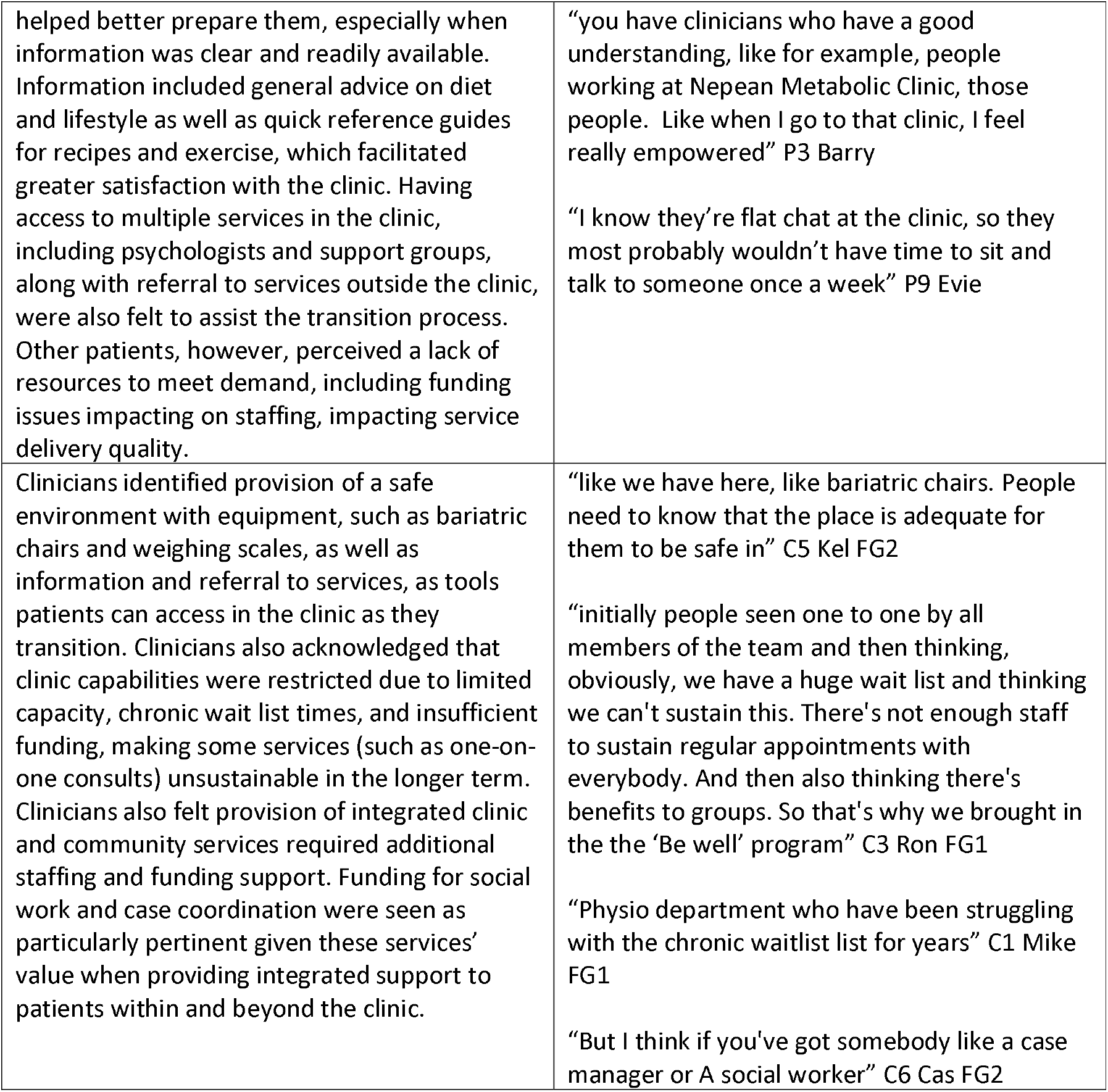

### 3.2 Individual-related factors influencing transition

Self-efficacy and self-promotion were individual-related factors which significantly influenced transition, with patients able to address their own support needs resulting in more successful transitions. Conversely, dependency on the clinic and its services, the presence of multiple comorbidities, as well as social issues and barriers, such as mental health problems and life-long weight issues, were likely to negatively impact the transition experience (Table 3).

**Table 3:**

| **Individual related factors influencing transition** |  |
| --- | --- |
| **Sub-themes** | **Excerpt** |
| **Self-efficacy and self-promotion**<br>Self-efficacy, motivation, and mindset were apparent in influencing transition experience. Patients who had a positive transition experience or were in the process of preparing their transition reported greater self-efficacy. This was observed in the language patients used to describe how they were supported through learning, self-motivation and setting up their own support networks. Patients also identified the importance of motivation and receiving encouragement in continuing to progress beyond the clinic. This motivation | “I don’t know if I need any support [from the clinic] at this point...I think, for me, I've done a lot of reading and trying to understand how my body reacts with different foods” P16 Jules<br>“I did not take the surgery, because I thought no. After the surgery I have to follow that lifestyle. If I can do it after the surgery, why can’t I do it now?” P9 Fran<br>“But I know it's their encouragement has sort of taught me how to do how to know what's right” P23 Tess |
| could come from either themselves (health-conscious mindset) or from others (i.e., peers, clinicians or family).<br>Not surprisingly, the converse created issues for other patients, who reported a lack of motivation hindered their transition, as it was difficult to maintain and continue progress, and achieve goals when feeling 'lazy'. | "we all, not that we're health conscious because we broke the rules quite often, but we'd all be saying, oh, you know, maybe that's not a good choice. And, you know, if we do want a snack, well, then it's Hummus, then it's healthy, healthy dips and stuff. We try, even if we don't always get it right." P23 Tess<br>"I always get these plan - you get these plans in your mind...then I just don't follow through with them. I procrastinate a lot" P2 Adrian |
| Clinicians agreed that patients with greater self-efficacy and ability to problem solve were more likely to successfully transition, as they were able to address their own support needs by leveraging existing support to form support networks and identify how to manage their health issues. Clinicians felt patients with low self-efficacy had difficulty accessing support on their own, as well as motivating and supporting themselves when encountering difficulties. | "focused on bringing in the supports into their world as they're transitioning out...So it's sort of wrapping the community around them to try and support them as they carry on" C2 Pat FG1<br>"language that sounds reassuring, i.e. I realize what I need to do now, which is address all these other things." C7 Pip FG2<br>"And here are some examples of networks that they can make, but that they physically needed to go out and. Yeah, hold their hand while they were doing that." C5 Kel FG2<br>"more variable in terms of capacity to take stuff on or willingness to take stuff on so well that sort of individualized characteristic" C2 Pat FG1 |
| **Dependency, comorbidities, and social issues**<br>Having a codependent relationship with the clinic was a significant barrier to transition. Several patients reported that they relied too heavily on the service and felt unable to make progress after they left. As a result, some patients felt unsupported by the clinic and did not like the reduced contact during transition, which resulted in dissatisfaction and negative perceptions of transition.<br>Patients also reported that comorbid conditions and social issues often hindered a successful transition. Some patients said these factors impacted on their ability to maintain exercise, attend clinic appointments and participate in activities, leading to an inability to maintain or lose more weight. Furthermore, almost all patients reporting mental health issues as a barrier to transition, as they impacted on their ability to participate in and complete tasks, including self-care and social activities. Life-long weight issues were an added complexity | "I'd want to be put back to the clinic. I lost 20 kilos like that, and I gained it like that...and when that come up at Christmas time about them [the clinic] cutting us, you can ask any of the girls, I was hysterical. I was in tears" Keira Kingswood Focus Group<br>"they [the clinic] had the oomph to say, yes, they'll keep up supported and all that sort of, but then over the time it just got slack" Rosebud Kingswood<br>"I get back on the bike and I'm fine and I'm going along and all of a sudden something else has happened and I can't use it. I feel like I'm taking once step forward and two steps back" P1 Anna<br>"liver issue doesn't help with not losing weight. And my pre-existing thyroid condition" P23 Mel<br>"I'm kind of really neglecting myself. Not taking |
| around transition, given the ease of falling back into deep-rooted patterns of behaviour and lifestyle. | care of myself some days. I don't clean anything. That's not me. And the more and more things start to build up in the home, the more I get stressed" P14 Jane<br>"because you've got to understand that for most people, they've been living with this obesity for a long time. So, it's easy to go backwards" P3 Barry |
| Clinicians identified codependency as a significant barrier to transition, describing some patients as being overly 'needy', and being unwilling to transition to independence. Clinicians felt that these patients resisted transition by over relying on the clinic and clinicians, despite being in the process of transition, and expressed displeasure when support was weaned.<br>Clinicians also found patient complexity level in terms of presence of comorbidities, mental health conditions, and social personal issues made a difficult transition more likely than in patients with low complexity. | "So I think a lot of people don't know who to go to for support and they hang on to us" C6 Cas FG2<br>"they would still continue to ring the service every week or email or be in touch with clinicians on a weekly basis" C3 Ron FG1<br>"became very reliant on each other and were very upset that we had to drop...to monthly, that didn't go down well!" C3 Ron FG1<br>"someone who's got less of those complex issues that are sort of impacting on their experience, here, then you've actually got a little bit more wiggle room to manage with some of those other issues" C2 Pat FG1<br>"So whereas when I think there's when there's a huge number of other chronic care issues, a lot of family disruption and social disruption limited skill levels in terms of engagement with the world, I think that would be sort of where things are a little less functioning" C2 Pat FG1 |

### 3.3 Bridging the gap to facilitate transition

Several factors were identified as being necessary to bridge the gap to facilitate successful transition out of the clinic. This included the need for transitional and ongoing integrated community care, through the provision of professional supports and services, such as general practitioners, allied health, and social services. Community support structures, including peer support groups were also identified as a need. These support groups were deemed as being particularly important as patients were frequently socially isolated. Several guiding principles underpinning how these groups could be successful were identified (Box 1). Additionally, during COVID-19 lockdowns, telehealth and online support were the only means to consult with patients. This allowed both the clinic and its patients to identify this as a component that could also facilitate transition (Table 4).

**Box 1.**
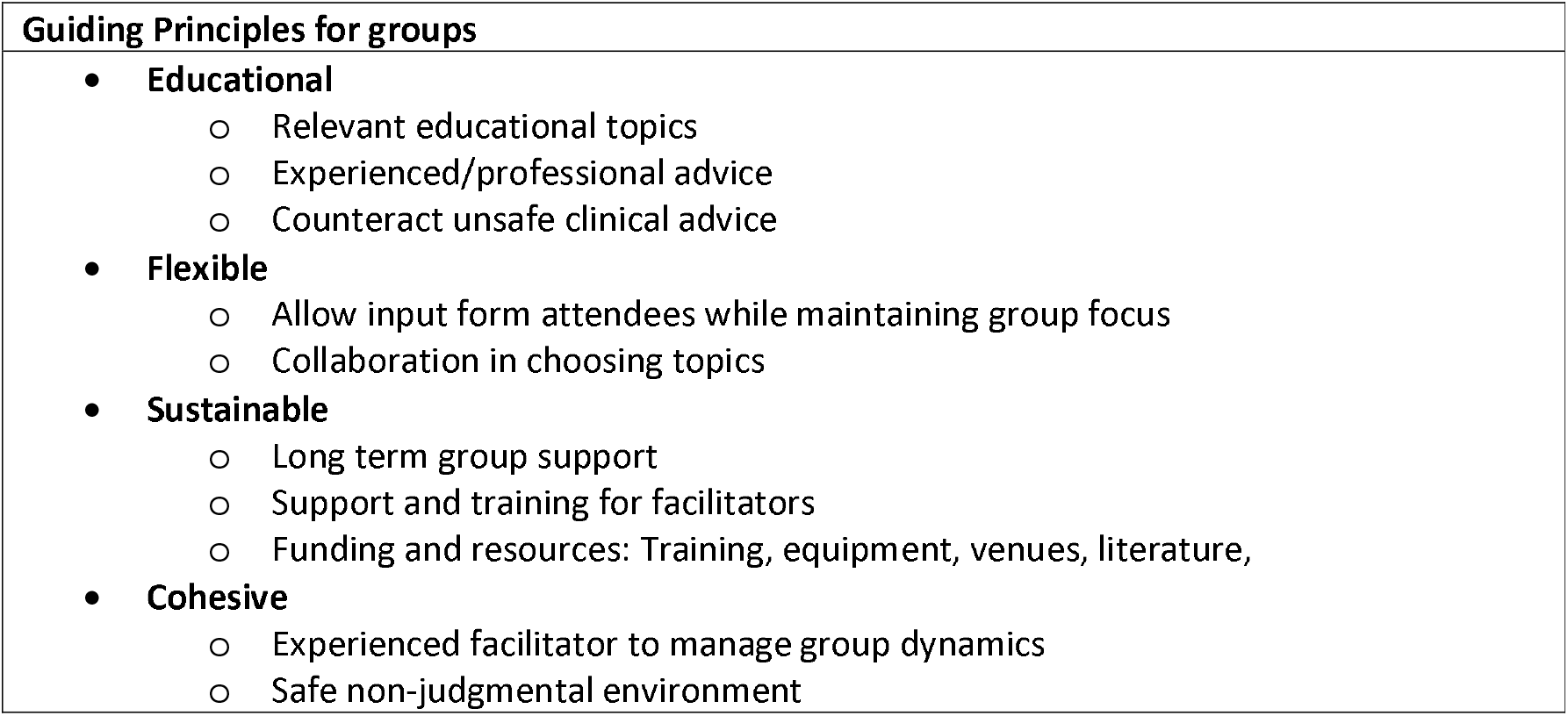
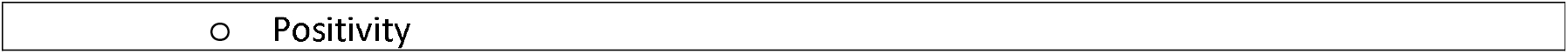

**Box 2.**
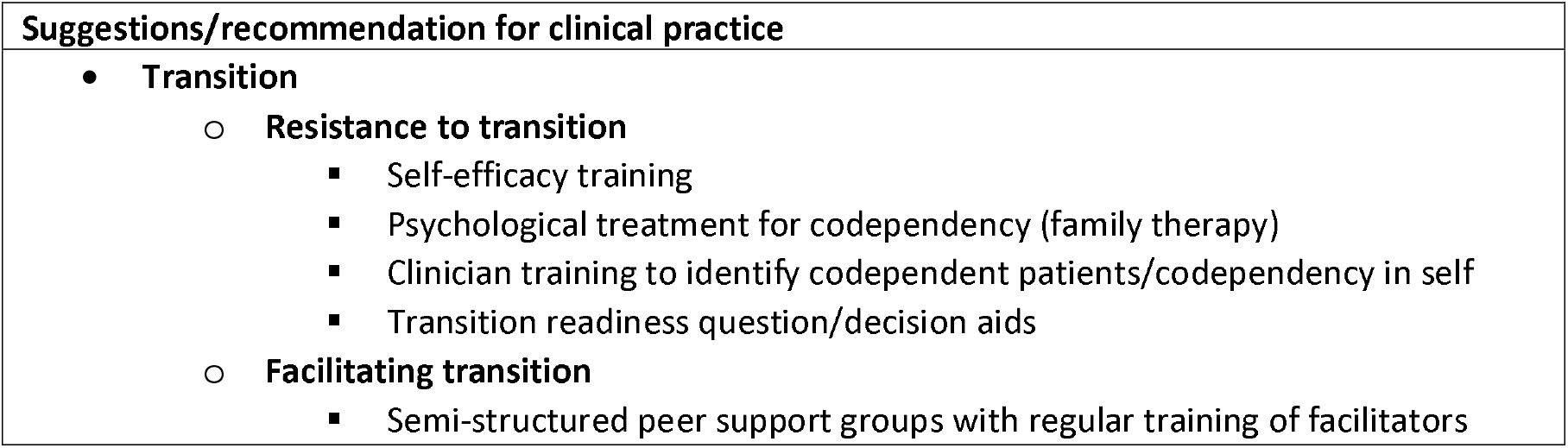
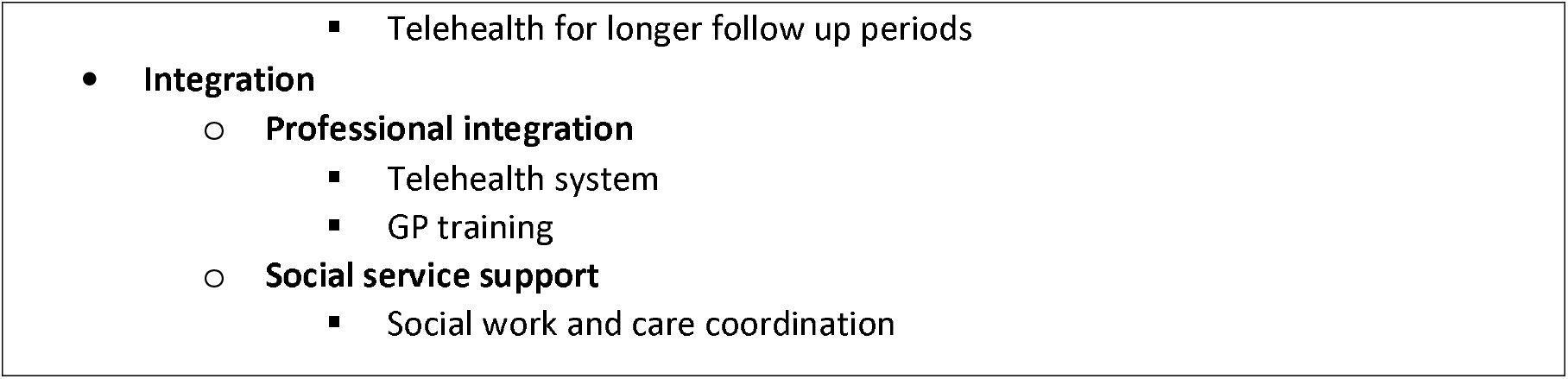

**Table 4:**
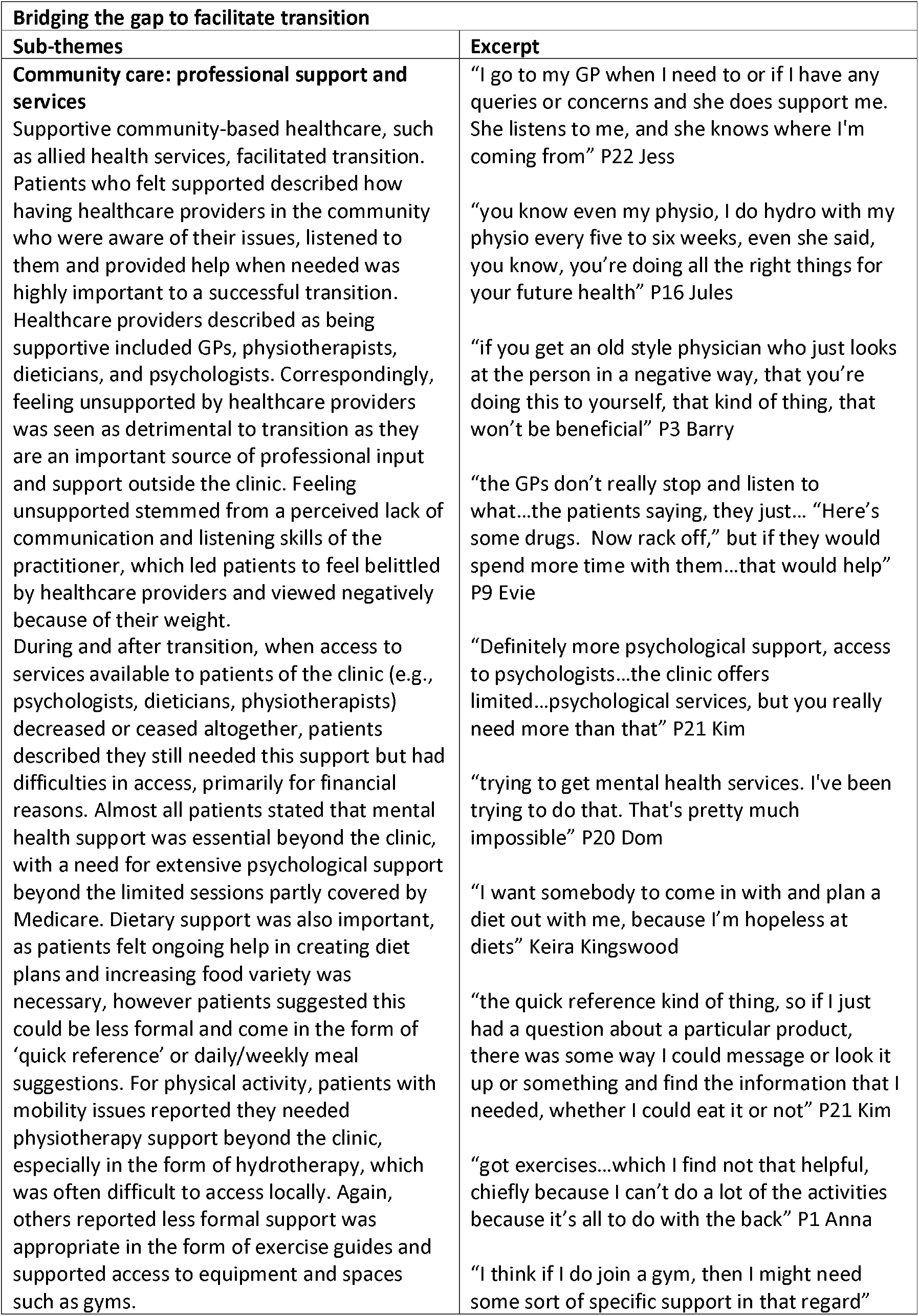
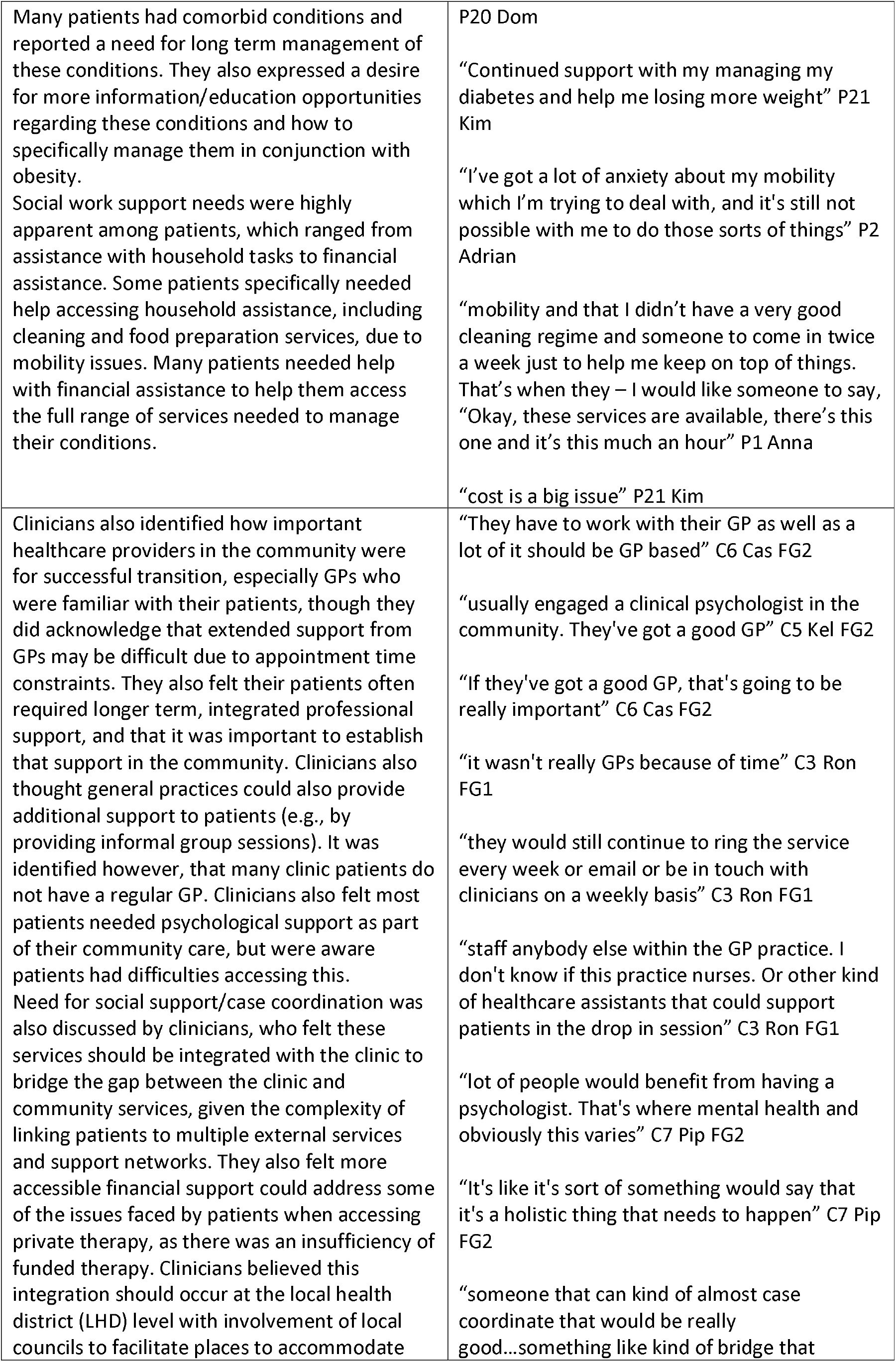
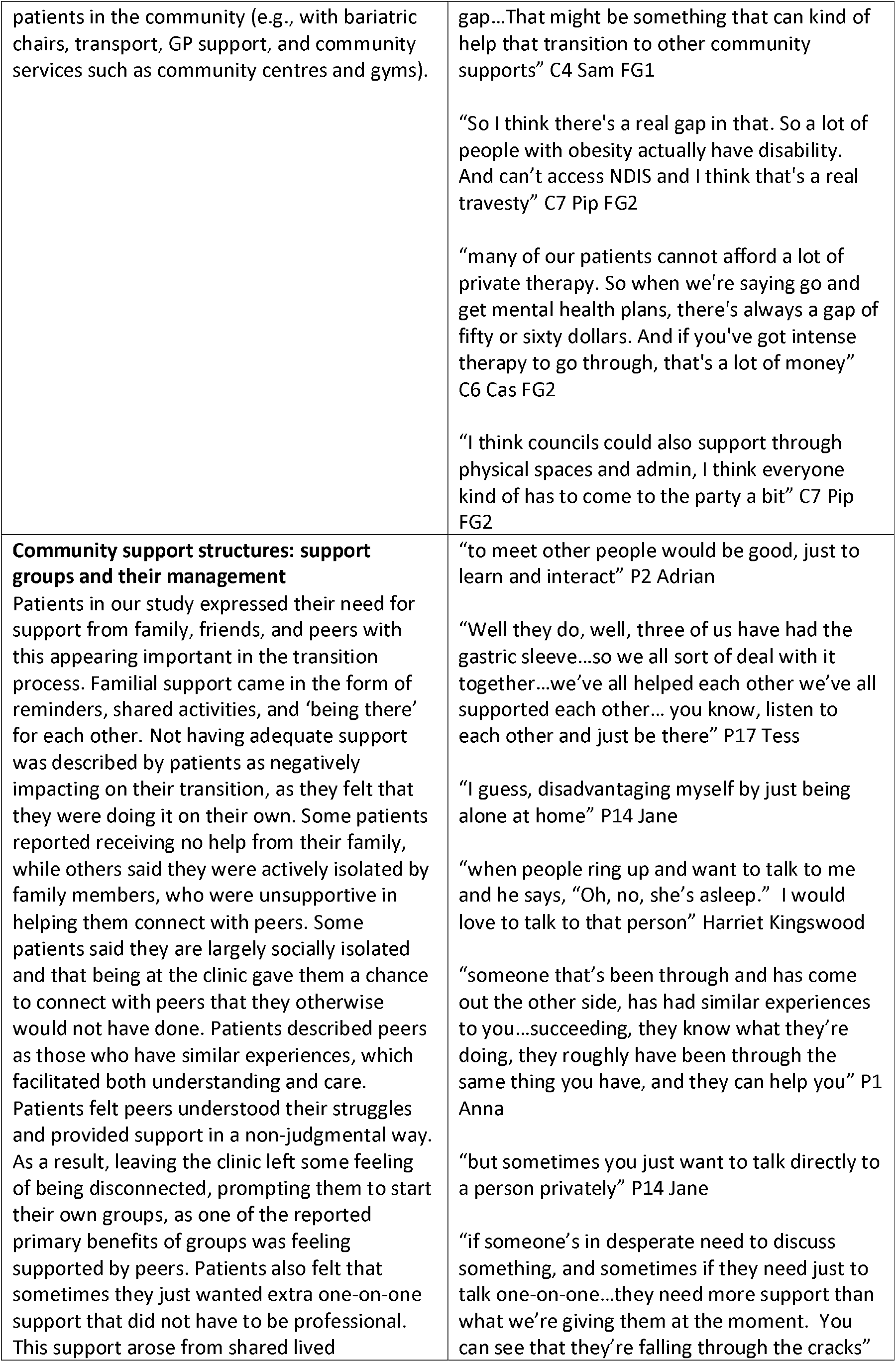
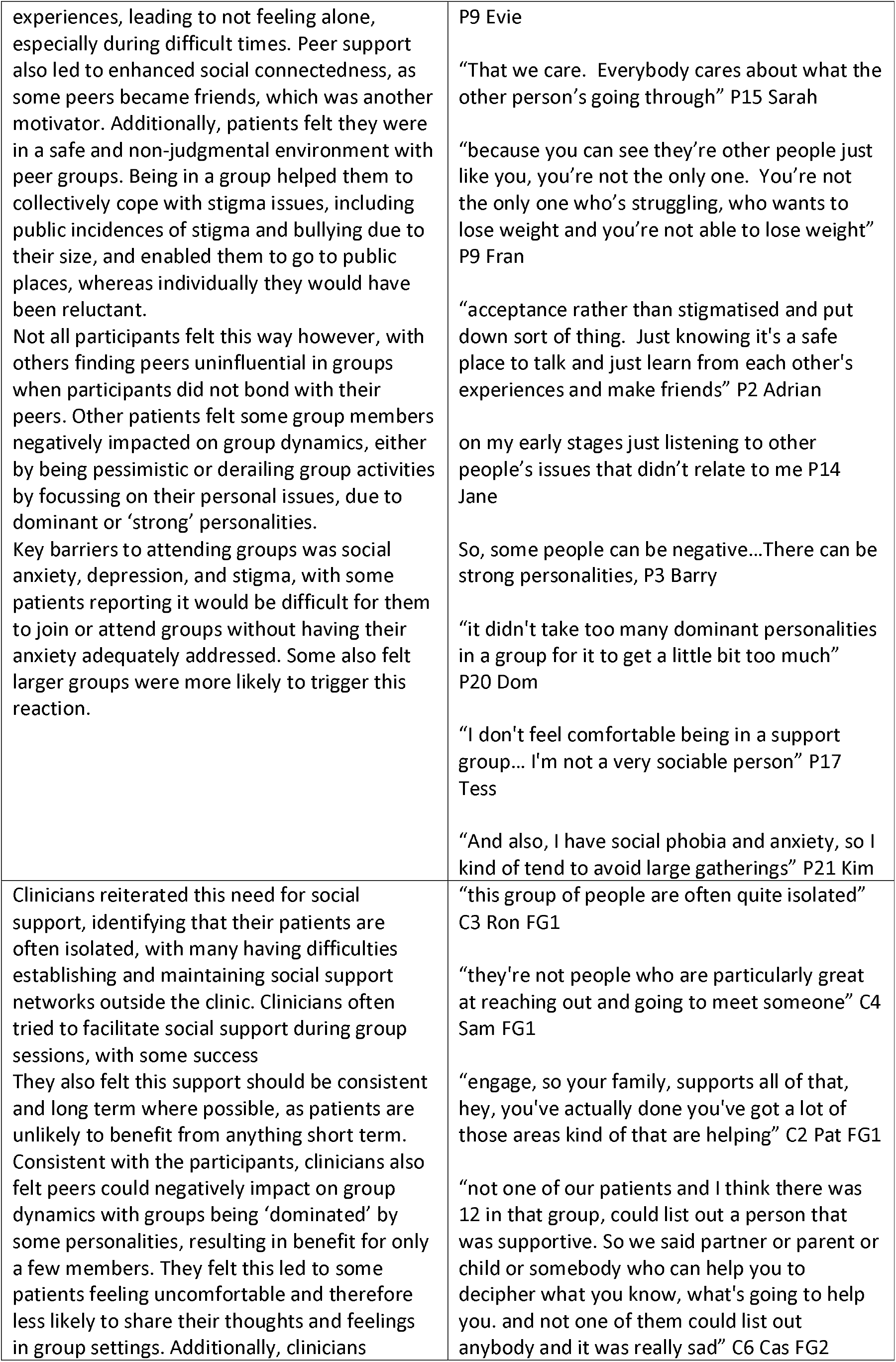
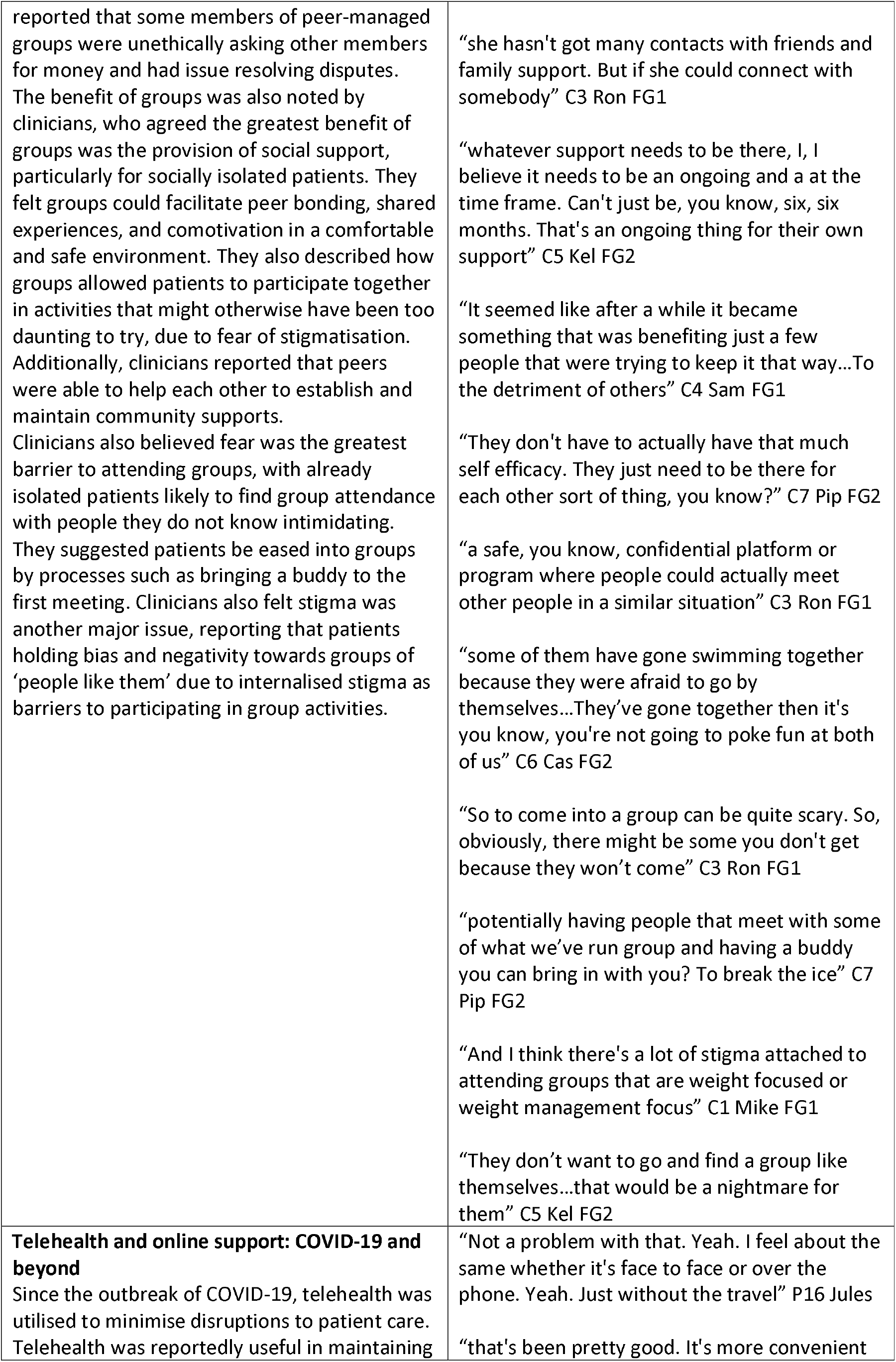
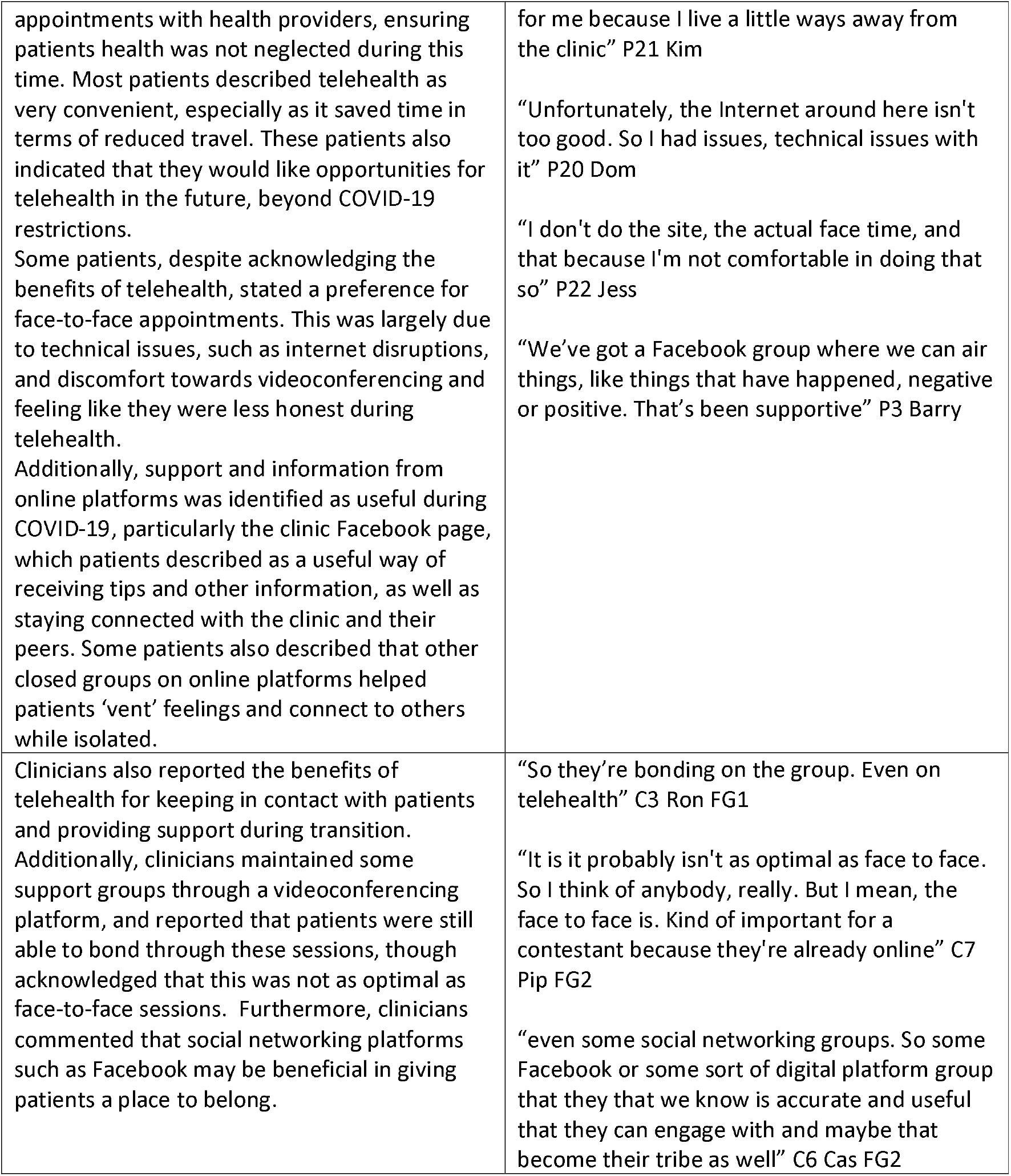

## 4. Discussion

Our results indicate that transition experiences vary considerably between participants who have attended a tertiary obesity service. While this appears to be a consequence of both individual characteristics and systemic factors such as transition process/policies, there is potential to facilitate a more congruent experience. This may be achieved by addressing factors such as patient dependency and self-efficacy, patient-provider communication, and transition processes. It was also apparent that the support needs of people living with obesity were only partly addressed during and following transition to community care. This is evident in the need for additional, integrated support from social and health networks that were identified by both the clinicians and patients in our study. Additionally, both patients and clinicians highlighted the difficulties in organising and coordinating these supports, suggesting a need for greater integration of services.

Dependency on the service and its clinicians was highly apparent. Level of dependency differed significantly between patients, and appeared to coincide with lower self-efficacy, with many patients lacking confidence to independently manage their own weight. These findings are consistent with a qualitative study of the follow up support of a weight management program for patients with obesity in the United Kingdom[11]. This study found an over-dependence on healthcare professionals hindered independent weight management, as patients deflected the responsibility of management to more authoritative figures, such as health professionals[11]. Lack of self-efficacy may be the driving force behind this dependence, suggesting some patients may need supports to boost their self-efficacy as an integral part of their transition process. Additionally, a sub-group of our patients tended to seek codependent relationships. This was apparent in our study among patients who expressed frustration and resentment for being dependent on family members, and for not being looked after enough by clinicians. While the evidence for this is limited, some studies have suggested that families with obesity display similar codependent characteristics to those seen among addiction conditions[12]. Such persons may also gravitate toward codependency with their clinicians, who can take on the role of ‘healer’ or problem solver while the patient takes on the role of passive recipient, with neither benefitting, as self-efficacy remains low. Resentment is also likely to ensue when patient problems are not ‘fixed’[13]. As such, patients are likely to benefit from access to greater support networks that may reduce reliance on professionals.

Codependency should be identified and treated in the context of low self-efficacy and self-esteem, anxiety, depression, lack of motivation, and isolation, which tend to cluster in both persons with codependence and obesity[14]. A primary aim of this strategy should be to increase self-efficacy. Family treatment may also be warranted in such a scenario, as it has been shown to be effective for mitigating codependence with other conditions, such as alcoholism and drug dependency[15, 16]. Further, clinicians may need to be more aware of their own propensity to enter codependent relationships with patients, hindering development of self-efficacy, while also causing burnout to themselves.

Patient-reported provider communication also influenced how patients felt about their transition. Patients who described a negative transition experience expressed lower satisfaction with provider communication, receiving limited information regarding transition and discharge, and limited opportunities for collaborative/patient-centred care. The opposite was true for those who reported positive experiences. These findings highlight the importance of clear communication between clinicians and patients regarding goals and expectations of treatment and discharge. The variation in patient experience suggests a need to identify dissatisfied patients early and to intervene to resolve communication issues to promote positive transition experiences. Patient-provider communication is crucial to patient satisfaction, and interventions to improve this have reported improved health outcomes, including reduced readmission in hospital settings[17]. It is important to note, however, that internalised weight bias is more strongly associated with perceived clinician bias than experienced weight stigma, which may account for the differences in communication reported by our patients, which ranged from ‘judged’ to ‘empowered’[18]. Patient dissatisfaction was more prevalent among those who were ‘resistant’ to transition, as such it may be beneficial to identify patients’ readiness for discharge/transition through standardised tools. This may be achieved through the use of decision aids and categorisation of patients to aid clinicians in identifying patients in need of additional intervention and referral to community care and support networks. While discharge readiness tools and decision aids are widely used in other care settings, there is limited evidence for their use in tertiary obesity treatment.

Our results strongly indicate the potential positive impact of peer support groups to facilitate transition. Many of the patients in our study described being socially isolated, a finding reiterated by clinicians. Social isolation can exacerbate mental health conditions, such as anxiety and depression, and reinforce internalised stigma[19]. This is bi-directional, as mental health conditions and stigmatisation can lead to greater social isolation[19]. The combination of these factors can lead to poorer health outcomes, especially during transitory phases of treatment. There is a potential for peer support groups to partly address these issues through social participation and networking in a supportive community environment. These groups have been recommended as being useful for mental health and chronic conditions, as increasing support networks are beneficial for mental health and can support health management behaviours[20]. Although efficacy of peer support groups varies between studies, support groups may be beneficial in weight loss and maintenance, especially as they foster a sense of community between participants[21]. While little is known regarding the usefulness of peer support groups to facilitate transition to community care, peer support currently forms part of transitional discharge models for transition from psychiatric hospitals to community, which have shown to be cost effective and result in lower readmission rates[22].

Additionally, peer support groups may help patients cope with stigma. The patients who took part in our study and their clinicians reported patients being more confident in participating in public activities, such as outdoor recreation, when in a group setting with their peers. Research suggests that support groups manage stigma by allowing persons to connect with a larger social support network, providing safe and non-judgemental environments, and allow for coping with confronting situations as a group, though these have not been evaluated in relation to obesity[23].

The structure and operation of peer support groups that the patients in our study were already part of varied considerably, making evaluation of effectiveness in this context difficult. As some groups were more successful than others, we asked patients and their clinicians to identify elements that may lead to more successful groups. These included the need for experienced/trained facilitators, while keeping groups less formal with the use of lay peer support workers rather than clinicians. Access to funding and resources to promote sustainability was also seen to be important, as well as ongoing training support for facilitators and collaboration with groups members on group structure and content, so as to mitigate the spread of misinformation and over reliance on group facilitators. These suggestions have been reported elsewhere, where facilitator training and experience have both been found to be vital to success of groups[24].

Barriers reported by patients to attending groups include depression, social anxiety, and stigma, while clinicians identified stigma as the primary barrier to attendance, especially in younger patients. It has been documented that mental health conditions and stigma negatively impact social participation and this correlates with findings that obesity related reduction in social participation is more common in younger individuals and appears to decline with age[25]. While stigma has not been explicitly reported as a barrier to support group attendance, research suggests that people with obesity hold strong explicit and implicit anti-fat attitudes and hence may not want to associate with others with obesity[26]. Clinicians in our study suggested that patients should be eased into groups through processes such as bringing a buddy to their first meeting, though little evidence exists for methods to address participation impacted by stigma. Further research is needed to explore methods to increase participation in groups impacted by stigma.

Telehealth may also be a potential ‘outreach’ support for patients as they transition out of the clinic. Telehealth was utilised by patients and clinicians due to lockdowns during the COVID-19 pandemic. Most patients found telehealth to be a convenient way of connecting with clinicians, especially as it addressed barriers such as transportation and time constraints. Literature suggests telehealth as an effective mode of health management for patients, especially in providing aftercare, though limitations to its use need to be addressed[27, 28]. These include access to adequate internet and related equipment, such as microphone and/or webcam, and telecommunication devices, and digital literacy[28]. There is potential for the application of telehealth in transitioning patients out of tertiary services to community care, by allowing for longer follow-up periods while maintaining clinic efficiency, which is especially useful for more complex patients[27]. Telehealth also has the potential to be part of an integrated care framework, by facilitating transition to community care as well as communication and training between tertiary and primary healthcare practitioners[29].

The New South Wales integrated care strategy aims to relieve the burden from individuals in accessing care through the integration and coordination of services that enable seamless transitions of care, especially between tertiary/secondary services and community care[30]. Our findings indicate pressure on patients and clinicians to establish and access care networks in the community, leading to a lack of adequate integration. Both patients and clinicians reported that following transition to the community, patients continued to require dietary, mental health, and physiotherapy support, but some had difficulties in setting these up. Clinicians also identified that handover to general practitioners (GPs) were difficult, as not all patients had regular GPs and not all GP practices could provide adequate support to patients. These findings are concurrent with research indicating GPs are best situated to provide long term community care for people with obesity but require additional training, support, and role clarification to achieve this[31-33]. This is particularly important, as it has been identified that GPsare ill equipped to manage obesity with the current level of training and tools at their disposal. However, interventions to increase GP confidence and self-efficacy can mitigate this[34, 35]. Additionally, this training should address the weight bias of GPs, which is extensively reported in the literature and deters patients from seeking their care[34]. Patients should also ideally be able to access multidisciplinary teams through primary care, which would help address their needs post-transition, though the lack of recognition of obesity as a chronic disease in Australia and associated lack of funding limits this option[2]. Considering the pressure on public health services to cope with rising obesity rates, integrated care networks may relieve some of this burden by addressing fragmented care, often leading to poorer outcomes and greater costs[3, 4].

The patients also identified a need for home care, transportation services in the community, and specialised equipment, a finding reported when transitioning people with obesity from hospital to home[36]. Clinicians also reported that patients often required disability support for physical and mental conditions but had difficulties in access and that patients were unlikely to find equipment tailored for them outside of the clinic. This is in accordance with current literature which shows that the cooccurrence of obesity and disability is increasing[37]. In addition to this, extreme obesity on its own impacts functionality and patients often need assistance with activities of daily living (ADLs), as well as specialised equipment such as bariatric chairs and pressure reduction mattresses, which many be costly if not reimbursed[36]. These findings suggest a need for the examination of the potential for broader social service support for people with obesity. Extreme obesity is not recognised as a disabling condition and is not eligible for the Australian National Disability Insurance Scheme (NDIS), which provides information, reimbursement, and connections to community service for people with disability[38]. As such, patients require alternate processes to address their social service support needs, due to this lack of recognition.

Additionally, patients in this study reported they did not have access to weighing scales that could accommodate their weight outside of the clinic. This negatively impacted their ability to monitor their weight in a home/community setting. Studies on access to weighing scales are limited, though it has been reported that most commercial scales have a maximum weight of 150-180 kgs, and that scales with maximum weight above this range were more costly and less common[39]. As weight monitoring can be beneficial to weight management, patients are disadvantaged in this regard due to reduced access to weighing scales[39]. Lack of access to weighing scales, and social service support needs including specialised equipment, may require the integration and establishment of additional support networks which can be managed/addressed through social work and care coordination as well as from services such as the NDIS, through recognition of the impacts of obesity regarding disability.

The social service support needs of people with obesity may be addressed through social work and care coordination, which are vital to receiving integrated care as they address the community support needs of patients. The role or social work in obesity is becoming more apparent. It has been recently argued that obesity is increasingly recognised as a social justice issue, due to social determinants and weight discrimination associated with obesity, and hence a need for social work intervention[40]. Consequently, it is within the scope of social work practice to mobilise services and supports to address socioeconomic and psychosocial needs of people with obesity[40]. Care coordination also plays a significant role, as care coordinators can assist in navigating healthcare systems through support, information, and connections to other services, which can be beneficial for patients with obesity, due to their complex needs. A review of systematic reviews on care coordination interventions found improved health outcomes in a range of conditions including glycaemic control in patients with diabetes, service continuity in patients with mental illness (e.g., including lower depression severity and improved adherence), reduced hospital readmissions, and improved mortality in patients with heart failure and stroke[41]. Further research is needed on the potential for care coordination for people with obesity in tertiary health services, especially as it may address reliance on these services and serve as a bridge to facilitate integration with community services.

Our study was limited by the lack of heterogeneity of the sample, as most participants were female and above the age of 45, though this reflects the demographics of patients attending the service. Additionally, while our results may be transferable to other settings, they are not generalisable. However, this is not the purpose of qualitative research, which rather aims to deeply explore a particular topic to identify the important areas requiring future interrogation.

## 5 Conclusion

We aimed to explore the support needs of people living with obesity in an Australian healthcare setting as they transition from tertiary to community care through a qualitative exploration of the perceptions of patients and the clinicians involved in their care. It was found that service and individual factors, such as perceived provider communication and patient dependency, respectively, significantly influence transition experience. Additionally, patients require integration and enhancement of community support service and structure to address their transition needs. Future research should aim to address social community service needs following transition to ensure adequate community care that can support the maintenance of treatment outcomes.

## Data Availability

All data produced in the present study are available upon reasonable request to the authors

